# A Constructivist Grounded Theory Study Protocol: What works for whom? Therapists’ and adolescents’ perspectives on indication criteria for schema therapy

**DOI:** 10.64898/2026.05.14.26353229

**Authors:** M.H.E. Wilms, J. Roelofs, M.A. Alma, M.M. Rijkeboer

## Abstract

**Aim:** Schema Therapy (ST) is an evidence-based treatment for complex mental health problems rooted in early Adverse Childhood Experiences (ACEs). Although both individual and group formats have shown effectiveness, little is known about which format works best for whom. This question is particularly relevant for adolescents given their unique developmental needs. Despite over a decade of clinical experience with ST in adolescents, current research offers limited guidance on how to tailor the format to individual needs - resulting in a persistent gap between research and practice. This study aims to develop practice-based indication criteria for individual versus group schema therapy by integrating therapists’ expertise with experiences from adolescents’ who underwent ST.

**Methods:** This qualitative study employs a constructivist Grounded Theory approach. Data will be gathered through focus group discussions with schema therapists and individual interviews with adolescents. Therapists will be purposively selected based on experience with both therapy formats and with traumatized adolescents. Adolescents are eligible if they have experienced ACEs and have completed at least 20 sessions of ST.

**Results:** The analysis will result in a theoretical model that integrates therapists’ clinical reasoning and adolescents’ preferences.

**Conclusions:** This study integrates schema therapists’ expertise and adolescents’ lived experiences to develop actionable indication criteria for choosing between individual and group ST. By supporting informed clinical decision-making, the findings contribute to treatment personalization in adolescent ST and address key challenges such as suboptimal outcomes and treatment dropout. Moreover, the identified criteria provide a foundation for future quantitative validation.

**Authors summary:** MW, JR, MA and MR contributed to the conceptualization and design of the study. MW drafted the initial version of the manuscript. JR, MA and MR critically reviewed and provided substantial revisions to the manuscript. MW integrated all feedback and prepared the final version. All authors read and approved the final manuscript.

## Introduction

Adverse Childhood Experiences (ACEs), such as sexual abuse, domestic violence, and emotional neglect, are strongly associated with long-term psychological and behavioral problems (1). Individuals exposed to ACEs are three to six times more likely to engage in sexually risky behavior, experience mental health issues, and misuse alcohol. They are up to seven times more likely to self-harm, abuse drugs, or engage in interpersonal violence (2). However, timely and targeted psychological interventions may mitigate or even prevent these negative outcomes (3,4).

Schema therapy (ST) was specifically developed to treat the long-term consequences of childhood trauma and neglect (5). It is posited that ACEs, in interaction with a child’s temperament, lead to the development of early maladaptive schemas (EMSs): ingrained maladaptive mental representations of the self, others and the world. These EMSs can be triggered by specific stimuli, subsequently exerting a strong influence on emotional responses and cognitive processing. To manage the distress associated with activated schemas, individuals develop coping responses that manifest in schema modes - momentary emotional, cognitive, and behavioral states. Over time, EMSs and schema modes become more entrenched through maladaptive coping patterns, such as avoidance or proving the opposite, which may increase the risk of re-traumatization (6).

In treatment, ST aims to meet universal emotional needs - such as safety, autonomy, and spontaneity and play - that were unmet during childhood and contributed to the development of EMSs and schema modes. The therapy seeks to address these unmet needs through a range of interventions, including experiential techniques, limited reparenting, and empathic confrontation, applied in both individual and group formats (e.g., 5,7-10). Both treatment formats have demonstrated (cost-)effectiveness across diagnostic groups and age ranges, often with low dropout rates (e.g., 11-16).

Despite these promising outcomes, most studies focus on outcomes at the group level, leaving the question of which treatment format works best for whom largely unanswered. Individual and group therapy involve distinct therapeutic processes (17). These differences are especially relevant given the unique developmental challenges of adolescence. Compared to adults, adolescents exhibit heightened sensitivity to peer relationships, greater difficulty establishing trust, and more pronounced challenges in emotion regulation (18-23). Moreover, adolescents report varying preferences for each format (24,25). These developmental factors and preferences likely influence adolescents’ responses to different therapeutic contexts and underscore the importance of more nuanced treatment allocation.

The absence of clear guidelines for treatment allocation may lead to suboptimal matching between patients and therapy formats, potentially increasing dropout rates and diminishing overall treatment effectiveness (26-31). This issue is particularly pressing for adolescents: not only has the number of clinical trials involving children and adolescents declined over the past five decades (32), several studies also indicate that treatment outcomes in this age group are stagnating or even worsening (33-36). Increasing research efforts to understand what works for whom, and to personalize interventions for adolescents, may help close this growing treatment gap (36,37).

Practice-based evidence offers valuable insights into the personalization and context-responsiveness of interventions aimed at optimizing treatment outcomes (e.g., 38,39). Notably, ST has been applied in both individual and group formats to traumatized adolescents in clinical practice for over a decade. This sustained use suggests the presence of substantial practice-based knowledge among therapists, as well as rich lived experience among adolescents, both of which remain largely underexplored in the literature. This disconnection between research and clinical reality exemplifies the broader “practice–research gap” (40,41). Systematically capturing the perspectives of both therapists and adolescents may help bridge this gap by integrating clinical expertise with empirical evidence (38,42).

The primary aim of this study is to gain a deeper understanding of the factors that, according to both therapists and adolescents, influence the treatment selection process and contribute to treatment effectiveness in individual and group ST. By examining therapists’ clinical reasoning in choosing between individual and group ST, as well as the factors they associate with successful outcomes, alongside adolescents’ perceptions of and preferences for both formats and their views on what contributes to effective treatment. These insights will form a first step in the development of a practical, experience-based framework to support more personalized and effective care (43-45) and will serve as a foundation for future empirical validation.

## Study status

Data collection for this study is currently ongoing. Recruitment of participants has not yet been completed. No data analysis has been conducted and no analytic outputs or results are available. Recruitment will continue until theoretical sufficiency is reached, meaning the point at which additional data no longer contribute substantively to the development of emerging conceptual categories. Recruitment is currently expected to be completed by September 2026, although the actual endpoint will be determined by the attainment of theoretical sufficiency. Data analysis will commence thereafter, with initial analytic findings expected in January 2027.

## Methods and analysis

### Study design

This qualitative study employs a constructivist Grounded Theory (CGT) approach, as developed by Charmaz (46-48). CGT builds on the original Grounded Theory methodology by Glaser and Strauss (49), which emphasized data-driven analysis with minimal theoretical preconceptions. However, such a purely inductive stance is often unrealistic in contemporary research settings, where prior knowledge and institutional requirements (e.g., funding criteria) play a role.

CGT preserves the inductive and iterative nature of the original method, characterized by constant comparison and analytical flexibility, while also recognizing the co-construction of meaning between researchers and participants (Figure 1). It explicitly incorporates the researchers’ experiential knowledge, thereby enhancing the transparency, credibility, and practical relevance of the findings.

**Figure 1.**
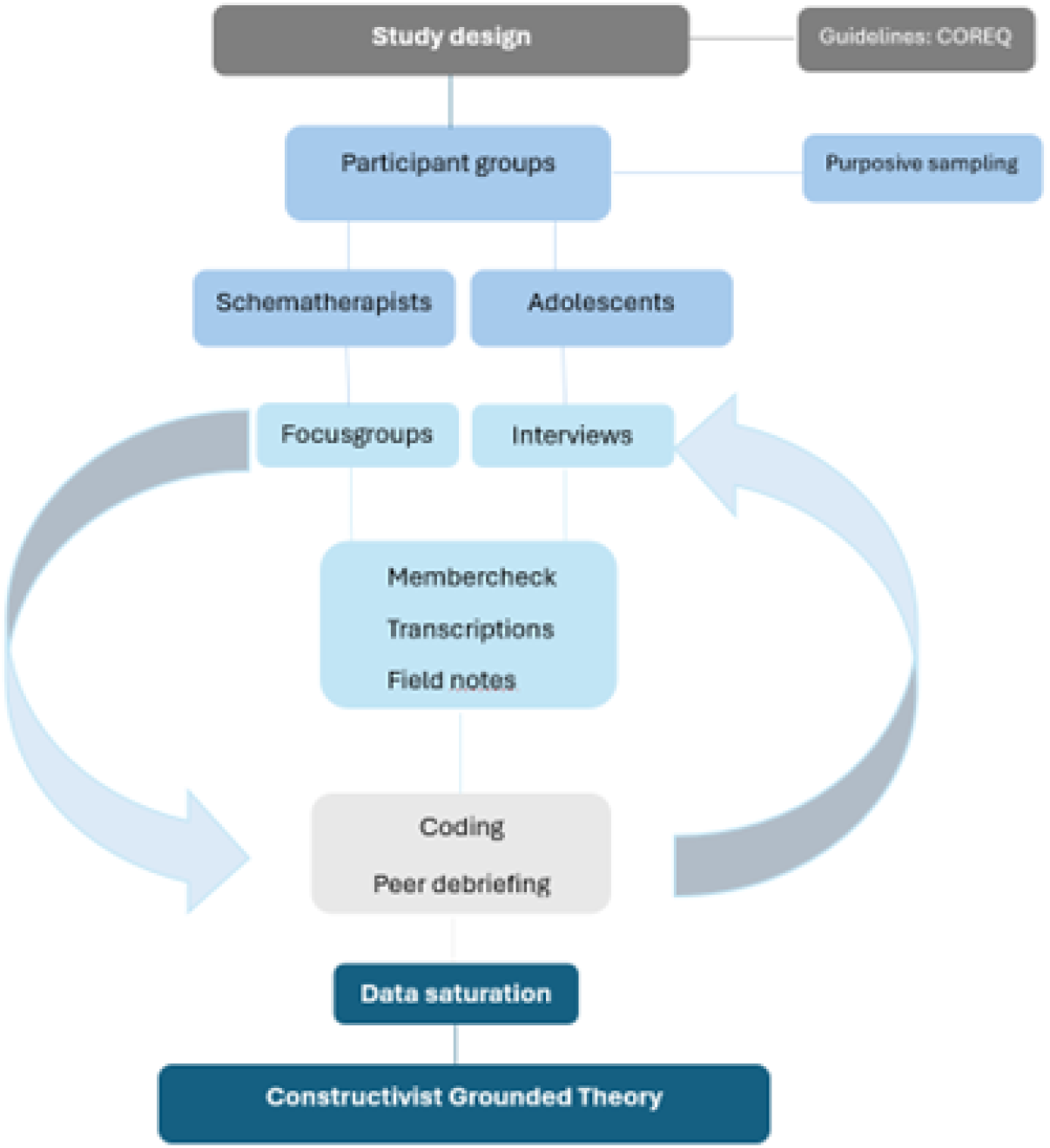
Flowchart of the study design

In line with this perspective, three of the four researchers in this study are experienced scientist-practitioners with specialized expertise in schema therapy, and two have extensive experience working with adolescents. Rather than being treated as a source of bias, this professional background is considered an asset that enriches the study’s interpretative depth and clinical applicability.

### Reporting guidelines

This protocol and the planned reporting of findings will follow the Consolidated Criteria for Reporting Qualitative Research checklist (COREQ; 50). COREQ was used prospectively during the protocol development and will be submitted as supplementary material.

### Participants and recruitment

This study includes two participant groups: one consisting of therapists and one consisting of adolescents. Therapists are eligible if they are certified senior schema therapists with experience in both individual and group ST and have provided ST to traumatized adolescents within the past five years. Adolescents are eligible if they have experienced one or more ACE(s), have received at least 20 sessions of individual or group ST between the ages of 16 and 23, and are able to reflect on and articulate their therapeutic experiences.

A purposive sampling strategy is employed to ensure the inclusion of participants with relevant experience and perspectives. All participants have no prior relationship with any of the research team members. Recruitment takes place via multiple channels, including mental health care institutions and social media platforms. Interested participants can register voluntarily by scanning a QR code on the recruitment flyer or by contacting the first author via email or text message. Upon registration, they receive an invitation letter, an information sheet, and a topic guide, each tailored to their respective group.

To support further recruitment, participants are invited to share the study information with others who may meet the inclusion criteria, enabling a form of snowball sampling.

## Data Collection

Data will be collected through focus groups and interviews conducted by MW. Focus groups, aimed at eliciting expert knowledge, will consist of three to eight schema therapists and last approximately ninety minutes. These sessions can be held either in person or online. Semi-structured interviews with adolescents will explore their personal experience with ST, last about one hour, and will be conducted in person, digitally, or by telephone according to the adolescent’s preference. No follow-up sessions are planned.

In line with the CGT, no formal pilot study will be conducted. Instead, the first interviews and focus group meetings will be used to iteratively refine the topic guides if needed. Participants will receive the topic guide in advance, outlining the open-ended questions that will be used. Data collection will be guided by Socratic questioning techniques to encourage reflection and depth. Guidance will be kept to a minimum, except when the discussion strays significantly off-topic, the conversation stalls, or key themes from the topic guide remain unaddressed.

There is a slight preference for group ST when working with adolescents, based on clinical experience and research with adults within the research team. To enhance data accuracy and minimize the risk of confirmation bias, several safeguards will be implemented: (1) the absence of predefined hypotheses; (2) regular team meetings and peer debriefings to support reflexivity and objectivity during analysis; (3) field notes will be taken during and immediately after each data collection session and (4) participant validation through member checking, whereby all participants will receive a summary of their sessions (Stahl & King, 2020).

All focus groups and interviews will be audio-recorded and transcribed verbatim and put in Atlas.ti (version 25; ATLAS.ti Scientific Software Development GmbH, 2024). Data collection will continue until data saturation is reached.

## Study status (methodological positioning)

To clarify the current stage of the study in relation to the analytical procedures described below, data collection is ongoing and recruitment has not yet been completed. No coding or formal data analysis has been conducted and no study results are available. This study is therefore still in the data collection phase.

## Data-analysis

This study employs triangulation across researchers, methods, and perspectives. Data will be collaboratively analyzed by the full research team, comprising three scientist-practitioners (MW, PhD candidate, female; MR, professor, PhD, female; JR, PhD, male) with expertise in ST, and one qualitative research specialist (MA, PhD, female). This diversity supports critical reflection, integrative interpretation, and enhances the credibility of the analysis.

Data will be analyzed using principles of CGT (48). No predefined themes will be applied; instead, the analysis will be guided inductively by the data and field notes. Initial coding will be performed by MW using ATLAS.ti, with regular peer debriefings to support reflexivity and refine emerging categories.

The analysis follows an iterative process to deepen understanding of emerging practice-based insights. Initial data will be explored to identify preliminary concepts and categories (*open coding*), which inform follow-up questions for subsequent focus groups and interviews. This cycle of data collection, coding, and refinement will be repeated multiple times. New data will be continuously integrated and compared with previous findings. The research team will meet regularly to review analytic progress and adjust the focus of data collection in response to developing insights.

Following open coding, a comparative analysis will be conducted across interviews and focus groups. Similar codes will be grouped, and broader patterns and categories - such as preliminary indication criteria, will be identified (*focused coding*). Based on these findings, theoretical sampling will be used to further explore and elaborate emerging constructs through additional data collection with therapists and adolescents. This process will continue until data saturation is reached.

The final outcome will be a theoretical model integrating therapists’ indication criteria and adolescents’ lived experiences in ST. To support interpretive validity, illustrative quotes will be presented in the results section.

## Discussion

This qualitative study aims to generate practice-based evidence regarding indication criteria for adolescents receiving individual versus group ST. By integrating expert knowledge from schema therapists with the lived experiences of adolescents themselves, it seeks to deepen our understanding of the key factors that guide optimal treatment allocation in this distinct developmental group. To the best of the authors’ knowledge, this is the first study to systematically explore practice-based indication criteria specifically for adolescent ST.

While previous research has established the efficacy of both individual and group ST across various diagnoses and age groups (e.g., 11-16), these studies primarily report on outcomes at the group level. Consequently, clinicians are often required to rely on experiential and tacit knowledge when determining the most suitable treatment format for individual adolescents. Within the context of early intervention psychiatry, where timely and appropriate treatment allocation is critical, the limited explicitation of such reasoning represents a notable gap.

In line with the growing emphasis on personalized psychological interventions (37), the present study is designed to make implicit clinical reasoning processes more explicit. However, it is important to acknowledge that the resulting indication framework will reflect perceived suitability and professional judgment, rather than empirically established predictors of treatment outcome. As such, the anticipated findings should be understood as descriptive and exploratory, offering insight into how clinicians and adolescents conceptualize treatment fit, rather than providing normative guidance on optimal allocation.

The use of a CGT approach (48) is well suited to these aims, as it allows theory to emerge from participants’ situated experiences and meaning-making processes. At the same time, this epistemological stance implies that the findings will be co-constructed and context-dependent. Therapists’ perspectives may be shaped by factors such as clinical experience, institutional setting, and familiarity with group-based ST, while adolescents’ accounts may be influenced by having already been allocated to a specific treatment format. These influences are inherent to qualitative inquiry and will be addressed through reflexive analysis and transparent reporting.

Consequently, the resulting indication framework should not be interpreted as a prescriptive or decision-making model, but as a conceptual tool to support clinical reflection and shared decision-making. By articulating how treatment suitability is currently understood in practice, this study aims to provide a foundation for hypothesis generation and subsequent quantitative research examining whether specific indication criteria are associated with engagement, outcomes, and optimal timing of individual versus group ST in adolescent populations.

## Implications and Conclusion

This study not only captures the expert knowledge of schema therapists and the lived experiences of adolescents, but also systematically translates these insights into concrete, actionable indication criteria to support clinical decision-making regarding individual versus group sST. By providing tailored guidance, this research meaningfully contributes to enhancing treatment personalization and may thereby addressing adolescents’ unique needs more effectively. The results are expected to have substantial clinical relevance and offer valuable insights to inform practice and guide further research in adolescent psychotherapy. This, in turn, has the potential to improve treatment outcomes and reduce dropout rates-two key challenges in adolescent mental health care.

Furthermore, the findings establish a robust foundation for subsequent quantitative research. Operationalizing the identified indication criteria in future studies will allow for their validation and refinement, ultimately strengthening the evidence base and clinical utility of ST.

## Strengths and limitations

A key strength of this study lies in the use of CGT, which allows for an in-depth exploration of complex clinical decision-making processes while remaining closely grounded in practice-based evidence. This methodological approach supports the systematic development of a practically applicable framework that reflects both professional expertise and contextual nuance. In addition, the study employs multiple recruitment strategies, including nationwide outreach, to enhance sample diversity and improve the representativeness of perspectives across clinical settings. Methodological rigor is further strengthened by the composition of the research team, which includes experienced scientist-practitioners with specialized expertise in ST and adolescent populations, alongside an experienced qualitative researcher. This combination supports reflexivity, analytic depth, and methodological consistency throughout the research process.

Despite these strengths, several limitations should be considered. Challenges inherent to participant recruitment and qualitative research - such as self-selection and potential sampling bias - remain difficult to fully eliminate, despite careful planning and strategic recruitment efforts. Consequently, some limitations in sample representativeness may persist, which may affect the transferability of the findings. Nevertheless, the results of this study provide a strong and conceptually robust foundation for future quantitative research, in which the identified indication criteria can be operationalized, tested, and further refined.

## Ethics and dissemination

This study underwent ethical review by the Ethics Review Committee Psychology and Neuroscience (ERCPN) at Maastricht University and was approved on 17th February 2025 under approval number ERCPN 288_117_11_2024, with approval granted for a period of three years. Findings will be disseminated through peer-reviewed journals, professional conferences, and clinical training events.

## Data Availability

No datasets were generated or analysed during the current study. All relevant data from this study will be made available upon study completion.

## Acknowledgements

The present study is supported by the Dutch “Stichting Achmea Slachtoffer en Samenleving (SASS)” with a grant.

